# An algorithm to detect dicrotic notch in arterial blood pressure and photoplethysmography waveforms using the iterative envelope mean method

**DOI:** 10.1101/2024.03.05.24303735

**Authors:** Ravi Pal, Akos Rudas, Sungsoo Kim, Jeffrey N. Chiang, Anna Braney, Maxime Cannesson

**Affiliations:** Department of Anesthesiology & Perioperative Medicine, University of California, Los Angeles, CA, USA; Department of Computational Medicine, University of California, Los Angeles, CA, USA; Institute of Sound and Vibration Research (ISVR), University of Southampton, Southampton, United Kingdom

**Keywords:** Arterial Blood Pressure (ABP) waveforms, Photoplethysmography (PPG) waveforms, Dicrotic notch (DN), Systolic phase duration (SPD), Iterative envelope mean (IEM) method

## Abstract

**Background and Objective:** Detection of the dicrotic notch (DN) within a cardiac cycle is essential for assessment of cardiac output, calculation of pulse wave velocity, estimation of left ventricular ejection time, and supporting feature-based machine learning models for noninvasive blood pressure estimation, and hypotension, or hypertension prediction. In this study, we present a new algorithm based on the iterative envelope mean (IEM) method to detect automatically the DN in arterial blood pressure (ABP) and photoplethysmography (PPG) waveforms.

**Methods:** The algorithm was evaluated on both ABP and PPG waveforms from a large perioperative dataset (MLORD dataset) comprising 17,327 patients. The analysis involved a total of 1,171,288 cardiac cycles for ABP waveforms and 3,424,975 cardiac cycles for PPG waveforms. To evaluate the algorithm’s performance, the systolic phase duration (SPD) was employed, which represents the duration from the onset of the systolic phase to the DN in the cardiac cycle. Correlation plots and regression analysis were used to compare the algorithm with an established DN detection technique (second derivative). The marking of the DN temporal location was carried out by an experienced researcher using the help of the ‘find_peaks’ function from the scipy PYTHON package, serving as a reference for the evaluation. The marking was visually validated by both an engineer and an anesthesiologist. The robustness of the algorithm was evaluated as the DN was made less visually distinct across signal-to-noise ratios (SNRs) ranging from -30 dB to -5 dB in both ABP and PPG waveforms.

**Results:** The correlation between SPD estimated by the algorithm and that marked by the researcher is strong for both ABP (*R*^2^(87343) =.99, *p*<.001) and PPG (*R*^2^(86764) =.98, *p*<.001) waveforms. The algorithm had a lower mean error of dicrotic notch detection (s): 0.0047 (0.0029) for ABP waveforms and 0.0046 (0.0029) for PPG waveforms, compared to 0.0693 (0.0770) for ABP and 0.0968 (0.0909) for PPG waveforms for the established 2^nd^ derivative method. The algorithm has high accuracy of DN detection for SNR of >= -9 dB for ABP waveforms and >= -12 dB for PPG waveforms indicating robust performance in detecting the DN when it is less visibly distinct.

**Conclusion:** Our proposed IEM-based algorithm can detect DN in both ABP and PPG waveforms with low computational cost, even in cases where it is not distinctly defined within a cardiac cycle of the waveform (‘DN-less signals’). The algorithm can potentially serve as a valuable, fast, and reliable tool for extracting features from ABP and PPG waveforms. It can be especially beneficial in medical applications where DN-based features, such as SPD, diastolic phase duration, and DN amplitude, play a significant role.

## 1. Introduction

In this paper we present a novel algorithm based on the iterative envelope mean (IEM) method [1] to detect the temporal location of dicrotic notch (DN) in arterial blood pressure (ABP) and photoplethysmography (PPG) waveforms. These waveforms are vital physiological parameters for health monitoring: ABP serves as a fundamental hemodynamic metric frequently used to guide therapeutic interventions especially in critically ill patients [2]; PPG, also known as the pulse oximetric wave, is a non-invasive technique used in anesthesia monitoring particularly for the assessment of blood oxygenation levels (SaO2) [3]. The presence of the DN in both ABP and PPG waveforms during the cardiac cycle holds valuable information about cardiovascular function and health. It is a small, downward deviation observed on the descending portion of these waveforms (see Fig. 1), occurring immediately after the systolic peak [4,5]. It is the result of the reflection of a wave at the aortic valve, following valve closure during the cardiac cycle [6,7,8, 9,10].

**Fig. 1.**
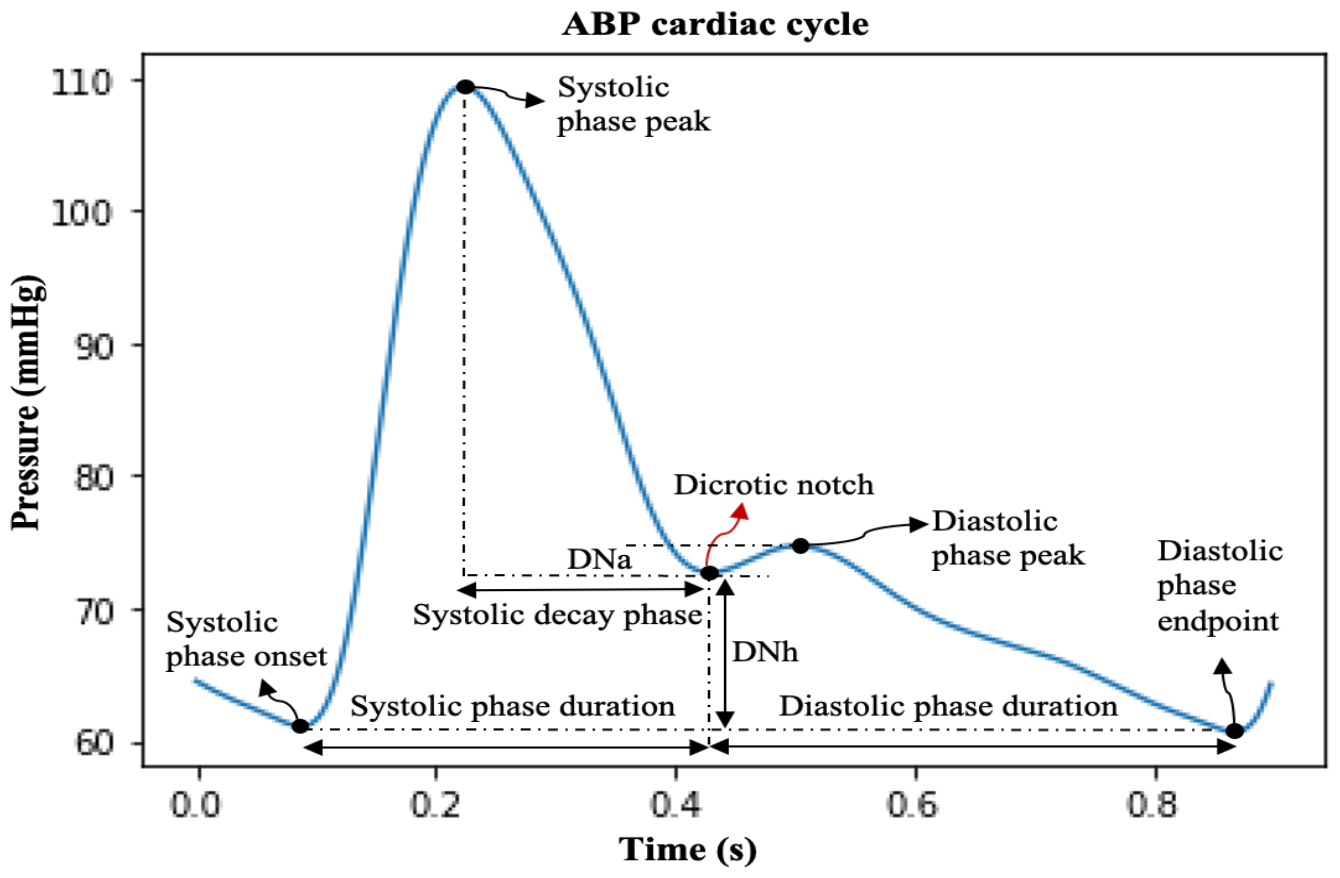
ABP cardiac cycle showing five key points: systolic phase onset, systolic phase peak (systolic pressure), dicrotic notch, diastolic phase peak, and diastolic phase endpoint (diastolic pressure); DNa: DN amplitude; DNh: DN height; systolic decay phase.

Due to its crucial role as a reference point in the timing of systolic and diastolic events, the DN has been used in numerous applications, including pulse wave velocity calculations [6,7,11], determining left ventricular ejection time [6,7], and models estimating cardiovascular function [7,12,13,14,15]. The DN also plays a crucial role in feature extraction from ABP and PPG waveforms by segmenting the cardiac cycles into distinct phases: the systolic phase (duration from systolic onset to DN), the diastolic phase (duration from DN to end of diastolic phase), and the systolic decay phase (duration from systolic peak to DN). These segmented phases/features may subsequently be utilized in feature-based machine learning models, contributing to a range of applications, including hypotension prediction [16] and non-invasive blood pressure estimation [17]. Furthermore, the positioning of the DN within the cardiac cycle has shown association with vascular tone [18,19,20]. Additionally, Abushouk et. al [4] underscores the clinical significance of several DN parameters: (a) DN amplitude (DNa): Higher DNa is associated with elevated peripheral vascular resistance, increased diastolic blood pressure, and the potential for weakened aortic valve closure, (b) DN height (DNh): Higher DNh signifies greater elasticity in the large arteries and effective aortic valve closure, (c) DN time duration or systolic phase duration (SPD): Longer SPD indicates robust systolic function of the heart. These applications emphasize the critical significance of accurate DN detection for enhancing clinical outcomes.

As mentioned in [7], pressure waveforms undergo shape changes as they propagate away from the heart due to variations in blood vessel properties and the influence of reflected waves. This can lead to the DN becoming less pronounced, sometimes visually undetectable, or just causing a slight inflexion in the curve [7]. As a result, identifying the DN becomes more challenging in distal pressure waveforms [7]. Furthermore, it is recognized that the shape of the DN tends to degrade with age in both ABP [7,21], and PPG waveforms [22,23]. Additionally, in PPG waveforms, the absence of a DN is associated with prevalent cardiovascular disease [24]. Therefore, it is crucial not only to identify the DN when it is clearly visible, but also to detect it within signals where it is substantially less apparent. These are referred to as ‘DN-less signals’. It is important to note that ‘detecting the DN’ means determining its temporal location within the cardiac cycle.

Many different algorithms have been developed to detect the DN [6,7,25,26,27,28,29,30]. However, either these methods were not thoroughly tested for noise robustness, or they were not examined for their ability to identify the DN in signals lacking a prominent notch (DN-less signals). Among these methods the 2^nd^ derivative method is the most widely used method for DN detection in the literature [28,31,32]. This approach employs the e-peak, the third peak within the 2^nd^ derivative of the PPG cardiac cycle, to estimate the DN within the cardiac cycle [31]. The primary limitation of the 2^nd^ derivative approach is its high sensitivity to noise. Various forms of noise including low and high frequency noise, motion artifacts, and baseline drift can distort ABP and PPG waveforms, posing significant challenges in accurately detecting the DN in these waveforms, often resulting in false detections or unreliable measurements. This highlights the need for the development of a more noise robust DN detection method.

In this paper, we present a new automatic DN detection algorithm based on the iterative envelope mean (IEM) method [1]. The IEM method divides a signal into two parts: the stationary and non-stationary contributions to the signal. The non-stationary output of the IEM method was used for detecting the DN. To evaluate its performance, we compare it with the 2^nd^ derivative approach for DN detection [31], which is extensively employed in the literature as the primary approach for DN detection.

The main contributions of this paper are:

1. a novel algorithm based on the IEM method to detect DN. This algorithm utilizes the non-stationary output of the IEM method to detect DN. The IEM method can detect DN in both ABP and PPG waveforms.
2. the algorithm has a low computational cost and the ability to detect DN not only in waveforms where it is clearly defined but also in waveforms with less pronounced DN characteristics.
3. evaluation of the performance of the algorithm using data from the MLORD dataset [33]. The results demonstrate that our algorithm outperforms the widely used 2^nd^ derivative method, highlighting its robust DN detection performance, and generalized applicability.

The rest of the paper is organized as follows. Section 2 describes the algorithm based on the IEM method. The dataset description and the performance evaluators are presented in Section 3. Sections 4 and 5 present the experimental results and discussion, respectively. Finally, we conclude our work in Section 6.

## 2. An automatic algorithm for dicrotic notch detection based on the iterative envelope mean method

The DN is the transition point between the systolic and diastolic phases of the cardiac cycle, as illustrated in Fig. 1. A block diagram of the proposed IEM method-based algorithm for DN detection is shown in Fig. 2. In this study, we processed 4-s windows of ABP and PPG waveforms using our algorithm. The waveforms are sampled at 256 Hz and the algorithm is implemented using the PYTHON programming language. Fig. 3(a) illustrates a 4-s window of the ABP waveform containing four complete cardiac cycles. The following sections describe the pre-processing and the IEM method stages of the algorithm.

**Fig. 2.**
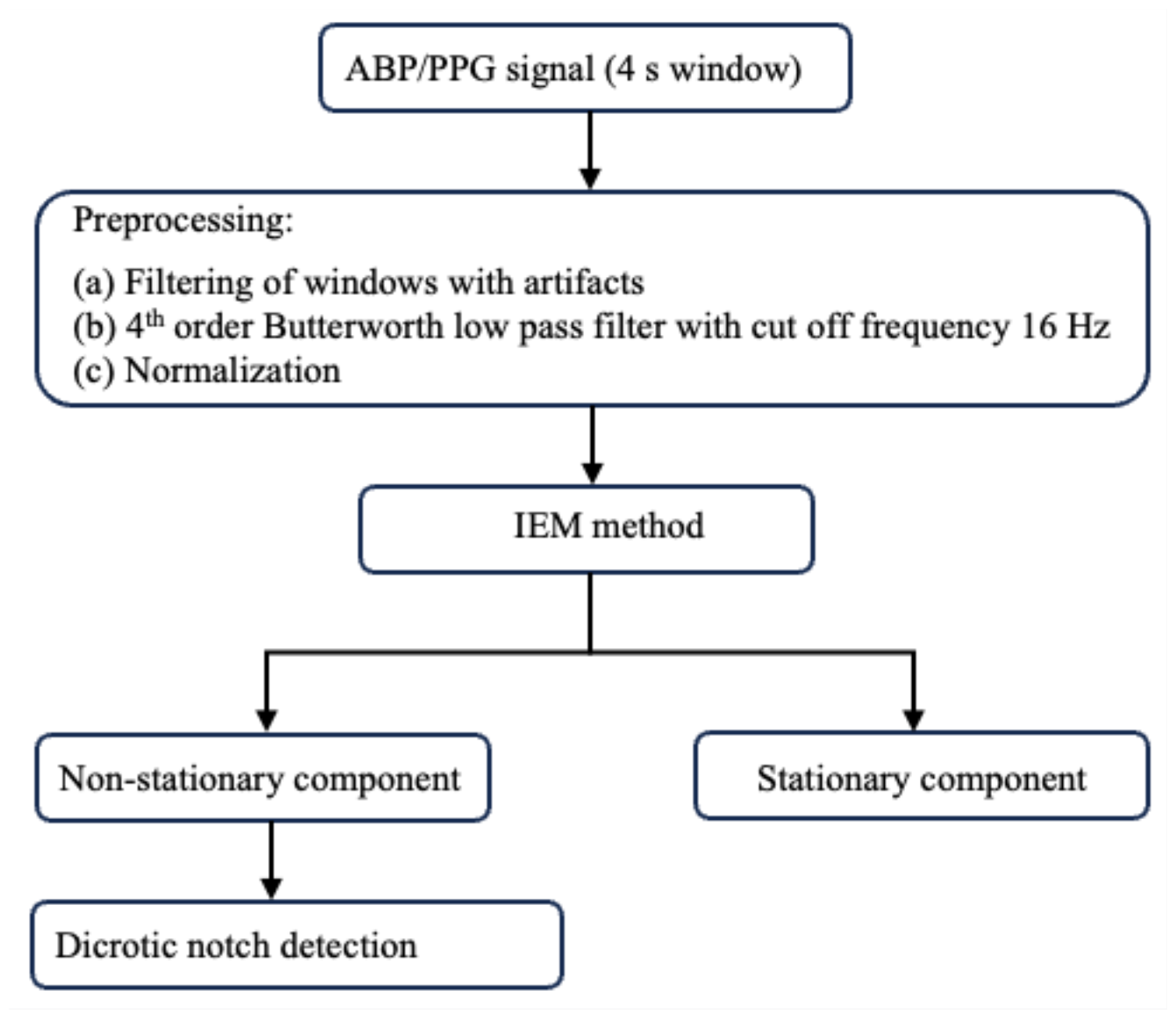
Block diagram of the algorithm.

**Fig. 3.**
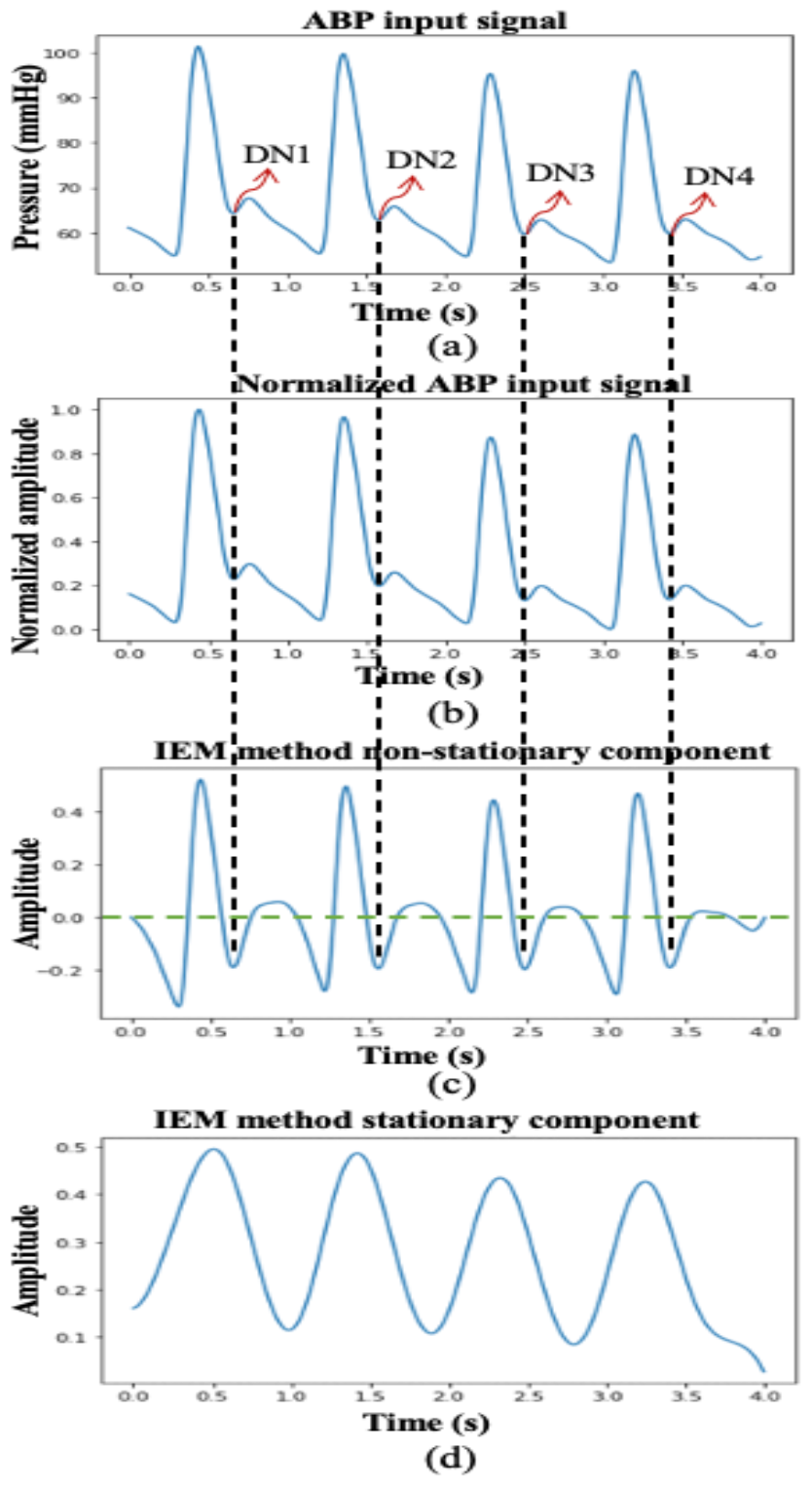
(a) ABP input signal with marked dicrotic notch locations by an experienced researcher; (b) Normalized ABP input signal; (c) IEM method non-stationary component; (d) IEM method stationary component.

### 2.1 Pre-processing

To eliminate windows with artifacts, we excluded any 4-s window that met one or more of the following criteria: containing zero or negative values, having three or fewer peaks exceeding 75^th^ percentile of the 4-s window amplitude, or having more than ten peaks. We used the ‘find_peaks’ function from the scipy PYTHON package for peak extraction. Secondly, each selected 4-s window was filtered using a 4^th^ order Butterworth low pass filter with cut off frequency of 16 Hz to remove high frequency noise [34]. Thirdly, the amplitude of the filtered and windowed signal was normalized using Eq. 1.

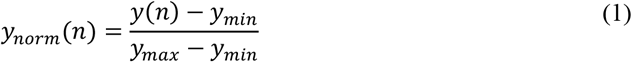

where *y(n*) represents the input signal, *n* is the sample index, *y*_*min*_ is the minimum value of the input signal within the window, *y*_*max*_ is its maximum value, and *y*_*norm*_ *(n*) denotes the normalized input signal. The normalized input signal is shown in Fig. 3 (b).

### 2.2 Iterative envelope mean method

The IEM method decomposes a signal into non-stationary and stationary parts [1]. The process involves smoothing the input signal. Finding the sample-by sample-mean of the upper and lower envelopes of the smoothed input signal and subtracting that mean from the original input signal. The resulting signal is then used as the input for subsequent iterations. After multiple iterations (*I* iterations), the IEM method provides an estimate of the non-stationary part of the original signal. Additionally, by summing the envelope means obtained from each iteration, it provides an estimate of the original signal’s stationary part. The IEM has previously been used in conjunction with the fractal dimension (IEM-FD) filter [1], for analyzing lung sounds. However, this study reports the first application of the IEM method for analysis of PPG and ABP waveforms to detect the DN.

The detailed working process of the IEM method for detecting the DN is as follows:

Step (1): The input signal is smoothed using the Savizky-Golay (SG) family filter and its first and second derivatives are calculated. The SG filter parameters are chosen following the guidelines proposed by Vannuccini et al. [35], with a polynomial fitting degree (*p*_*f*_) of 4 and a number of coefficients (*n*_*c*_) approximately one to two times the half-width of the shortest-duration feature of interest in the signal. In PPG and ABP cardiac cycle signals, the shortest duration may occur between any two consecutive key points within the cardiac cycle. The cardiac cycle consists of five key points: systolic phase onset, systolic phase peak, DN, diastolic phase peak, and diastolic phase endpoint, as displayed in Fig. 1. Since the duration of a single cardiac cycle is approximately 0.8 s, the duration between two consecutive key points will be approximately 0.2 s. In our dataset for which the sampling frequency is 256 HZ, the half width is approximately equal to 25 samples. The SG filter parameters used here are therefore *p*_*f*_ =4, *n*_*c*_ = 25 and order of derivation (*d*_*o*_) =0, 1 and 2 for smoothing the input signal, and for estimating the first and second derivatives of that smoothed signal, respectively.

Step (2): The locations of all the local extrema of the first derivative 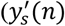 are identified and classified as either maxima or minima by examining the sign changes in the second derivative 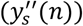 of the smoothed input signal *(y*_*s*_ *(n*)).

Step (3): The coordinates in the smoothed input signal, corresponding to the positions of each of the local maxima and minima in the first derivative, are calculated. Using cubic spline interpolation, the upper envelope (*UP*_*env*_*(n*)) is obtained by connecting the coordinates corresponding to local maxima of the first derivative, while the lower envelope (*LW*_*env*_*(n*)) is derived by connecting the coordinates associated with local minima of the first derivative. The envelope mean value is then calculated using the estimated upper and lower envelopes (Eq. 2)

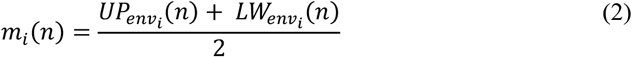

where *n* is the sample index in the input signal i.e. *n* =1, 2, …. *N* and *i* is the iteration number where *i*=1, 2,…, *I*.

Step (4): The computed envelope mean value is subtracted from the unfiltered version of the input signal to obtain the estimate of its non-stationary part (Eq. 3).

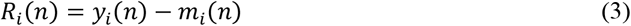

Step (5) If the resulting signal satisfies the stopping criterion shown in Eq. 4 it is taken to be the non-stationary part of the input signal otherwise, it is used as a new input signal: *y*_*i-*1_*(n*) = *R*_*i*_*(n*), and the process is repeated from step 1. The stopping criterion is given by:

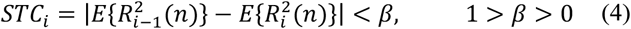

where E{.} denotes the expected value and has an initial value of *R*_*i-*1_ = 0. In this study we have used *β* = 0.1, and note that the IEM method employs the identical stopping criterion to that specified in [1,36,37,38].

The iteration process ends when the stopping criterion is met, at iteration *I*, and at this point the resulting signal is assigned as the non-stationary signal (NSTS) contribution to the input signal (Eq.5) while the summation of envelope means obtained across all iterations (*I*) is assigned as the stationary signal (STS) of the input signal (Eq.6).

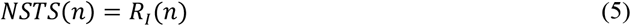

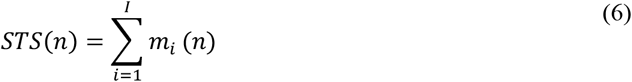

The IEM method non-stationary and stationary outputs are shown in Fig. 3(c) and (d), respectively. Although the non-stationary component of the IEM method can reveal the DN within a cardiac cycle, several further steps are required mark accurately the location in time of the DN. These involve identifying a valley within a cardiac cycle in the NSTS situated between the systolic peak and the diastolic endpoint (the end point of the cardiac cycle), as the DN location. As mentioned in [9], in addition to the DN, several other secondary waves are often generated by pressure reflections within the arterial system, which are frequently evident between the systolic peak and the diastolic endpoint in the ABP waveform, potentially introducing multiple valleys into each cardiac cycle within the NSTS. Moreover, non-physiological oscillations in both PPG and ABP waveforms can also contribute to the emergence of multiple valleys within the cardiac cycle. To address this issue, two conditions have been established: (1) the valley must be at least 0.1 s (25 samples) from the systolic peak; and: (2) the y-axis value corresponding to this valley in the NSTS must be less than zero. The first valley that satisfies both conditions is designated the DN location. The combination of the IEM method with the process of locating the DN within cardiac cycles in the NSTS is therefore collectively referred to as the IEM-based algorithm. In Fig. 3, the black vertical dashed lines represent the DN positions located within a cardiac cycle by the IEM-based algorithm, and the green horizontal line in the non-stationary component of the NSTS represents the zero-normalized pressure axis.

## 3. Analysis

In this section, we describe the dataset and the quantitative evaluators used for assessing the DN detection performance of the IEM-based algorithm.

### 3.1. Dataset

A large perioperative dataset (MLORD), comprising physiologic waveforms collected from 17,327 patients who underwent surgeries between 2019 and 2022 at the David Geffen School of Medicine at the University of California Los Angeles (IRB # 19-000354) [33], was utilized to assess the performance of the proposed algorithm. The MLORD dataset includes waveform data totaling over 72,264 hours in duration and 7.6 terabytes in size. This waveform data contains sampled digital physiological waveforms obtained from vital sign monitoring devices, such as Electrocardiogram (ECG) and PPG, as well as data recorded by other devices, including invasive ABP monitors. The data was sampled at 256 Hz. For our analyses, we utilized ABP and PPG waveforms only. A detailed description of the MLORD dataset may be found in [33].

In the MLORD dataset out of 17,327 patients, 4,901 patients have ABP waveforms and 17,170 patients have PPG waveforms. In this study, a total of 352,257 4-s windows from ABP waveforms, including 1,171,288 full cardiac cycles, and 1,100,689 4-s windows from PPG waveforms, representing 3,424,975 full cardiac cycles were used for the analysis. However, the marking of the temporal location of DN on all the cardiac cycles to serve as a reference is not feasible due to the dataset’s large size. Additionally, in certain instances, the DN or diastolic phase peak may be less distinct, especially in PPG waveforms, making it challenging to establish their temporal location as a reference. Therefore, to compare the performance of the algorithm with the existing 2^nd^ derivative method, we randomly selected 25,000 4-s windows from the ABP waveforms and the same number from the PPG waveforms where the DN could be observed within a cardiac cycle. After selecting the windows, an experienced researcher marked the temporal location of DN as well as the systolic phase onset within a cardiac cycle with the help of the ‘find_peaks’ function from the scipy PYTHON package. The first minimum before the systolic peak (largest peak in a cardiac cycle) was used as the onset point of the systolic phase. To ensure accuracy, the marking was validated by an engineer and an anesthesiologist. They conducted a visual examination of marked ABP and PPG windows. The selected ABP windows contained a total of 87,345 cardiac cycles; the PPG windows contained 86,766 cardiac cycles. To evaluate the performance of the IEM-based algorithm, the systolic phase duration (SPD) was used, which is defined as the time difference between the onset of the systolic phase and the occurrence of DN in the cardiac cycle [6]. The measured SPD, based on the marking, served as a reference. It is important to note that the systolic phase onset marked by the researcher is also used as the onset point for the SPD measured by the algorithm. Thus, any difference in SPD between the reference and the estimate from the algorithm is due to differences in the location identified as the DN only.

To evaluate the algorithm’s robustness in detecting the DN when it was less visibly distinct, we included all 352,257 4-s windows of ABP waveforms and 1,100,689 4-s windows of PPG waveforms. The algorithm’s robustness was tested across a range of signal-to-noise ratios (SNR) ranging from -30 dB to -5 dB in steps of 1 dB where ‘signal’ is defined as the root mean square amplitude of the non-stationary output of the IEM filter and ‘noise’ is its root mean square amplitude of the stationary output.

For the robustness analysis, to evaluate the performance of the algorithm, the reference SPD was calculated by initially decomposing each 4-s window into its non-stationary and stationary components. Subsequently, within each cardiac cycle in a 4-s window, we marked the temporal location of DN using the non-stationary component of the IEM method. We also employed the ‘find_peaks’ function from the scipy PYTHON package to identify the first minimum before the systolic peak (the largest peak in a cardiac cycle), which served as the onset point of the systolic phase. Using the marked DN and the identified systolic phase onset point, the reference SPD was calculated. Next, the stationary component of the IEM method was multiplied by a scaling factor and then added to the (un-scaled) non-stationary component. Scaling factors were chosen to give an SNR (ratio of non-stationary to scaled stationary component) ranging from -30 dB to -5 dB in steps of 1 dB. This had the effect of making the DN less visibly distinct. The IEM-based algorithm was then applied to this new signal and the temporal location of DN was detected. Using the detected DN after scaling and the systolic phase onset point calculated before scaling, the SPD for each sample was again calculated and compared with the reference SPD calculated before scaling to evaluate the robustness of the proposed algorithm. A block diagram of the analysis procedure in this study using the MLORD dataset is shown in Fig. 4 for better understanding.

**Fig. 4.**
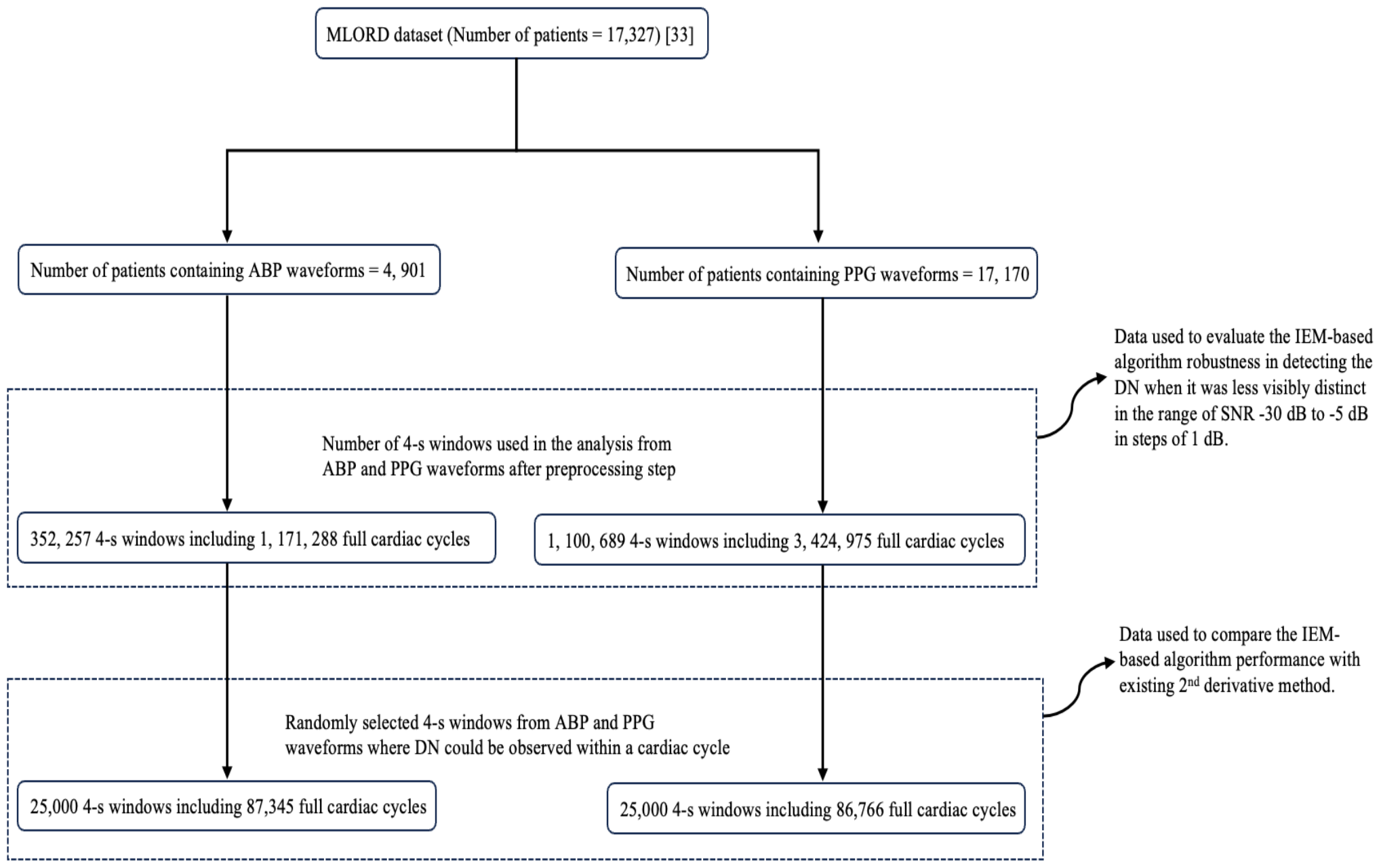
Block diagram of the analysis procedure using MLORD dataset.

### 3.2. Performance evaluators

The algorithm’s performance was assessed by comparing its estimates of SPD with those marked by an experienced researcher (as discussed in section 3.1). The evaluation was conducted using regression analysis [39], box plots [40], the rate of detectability (*D*_*R*_) [1,36], and the error of DN detection. Box plots were used to visually compare the difference between the reference SPD, as marked by the researcher, and the measured SPD obtained using the algorithm [1]. Additionally, comparisons were made between the proposed IEM-based algorithm and the existing 2^nd^ derivative method.

The closer the SPD difference is to zero, the greater the accuracy of the estimation method (algorithm or 2^nd^ derivative method) at collocating the DN with its position in the reference signal. The regression analysis demonstrates the strength of the linear association between the SPD measured by the algorithm and that measured by the researcher, The rate of detectability (*D*_*R*_) was used to assess the number of DNs detected by the algorithm, or by the 2^nd^ derivative method, within each 4-s window compared to the true number of DNs. We also calculated the error in DN detection, which is determined by the absolute value of the SPD difference.

In the context of robustness analysis, we used rate of detectability (*D*_*R*_) metric to measure the number of DNs within a 4-s window detected by the algorithm, after scaling compared to the number detected in the same window before scaling. We also computed error of DN detection using the difference in SPD before and after scaling at a given SNR. It is important to note that the systolic phase onset point detected by the algorithm for calculating SPD before scaling is also used as the onset point for calculating SPD after scaling. Thus, any difference in SPD between the before and after scaling is due to differences in the location identified as the DN only.

The process of calculating the rate of detectability, SPD difference, and the error of DN detection is as follows:

#### 3.2.1 Rate of detectability

The Rate of Detectability (*D*_*R*_) measures the ability of the IEM based algorithm to detect the DN within a cardiac cycle. *D*_*R*_ was calculated using (Eq. 7).

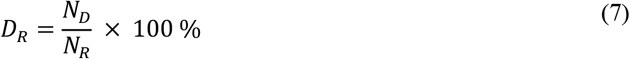

where *N*_*D*_ is the number of DNs detected by the proposed algorithm within a 4-s window and *N*_*R*_ is the number of reference DNs in the same window.

#### 3.2.2 Systolic phase duration (SPD) difference and error of dicrotic notch (DN) detection

The difference in SPD between the reference and the measured, or between before and after scaling was calculated using Eq. (8).

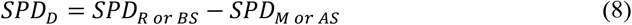

Where *SPD*_*R or BS*_ is the reference SPD or SPD before scaling in the context of robustness analysis and *SPD*_*M or AS*_ is the measured SPD by the algorithm or SPD after scaling in the context of robustness analysis.

Note that as mentioned earlier, any difference, in SPD between the reference and the estimate from the algorithm or in SPD between before and after scaling, is due to differences in the location identified as the DN only. Therefore, the absolute value of difference in SPD can be referred to as the error of DN detection (Eq. 9).

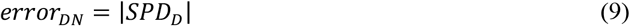

## 4. Experimental results

This section presents the results obtained from the IEM-based algorithm for DN detection in ABP and PPG waveforms, along with its performance comparison with the existing 2^nd^ derivative method and an analysis of its robustness in detecting the DN when it is less visibly distinct.

### 4.1 Performance of the IEM based algorithm in ABP and PPG waveforms

In Figs. 5 and 6, three different cases of ABP and PPG input signals are presented to demonstrate the algorithm’s performance. First, cases where the DN is clearly observable within a cardiac cycle are illustrated in Fig. 5(i) and Fig. 6(i) for ABP and PPG inputs, respectively. Plots labeled as (a) in these figures depict the input signal window, each lasting 4 s, with DN locations marked by an experienced researcher. Following this, cases where DN is less evident within a cardiac cycle are shown in Fig. 5(ii) and Fig. 6(ii) for ABP and PPG inputs, respectively. Plots labeled as (a) in these figures also display the input signal windows with marked DN locations, providing a reference for validation. Lastly, cases where DN is absent within a cardiac cycle, referred to as DN-less signals, are presented in Fig. 5(iii) and Fig. 6(iii) for ABP and PPG inputs, respectively. In Figs. 5 and 6, plots labeled as (b) depict the curves for the normalized input signal, plots labeled as (c) show the curves for the non-stationary component of the IEM method, and plots labeled as (d) display the curves for the stationary component of the IEM method. Importantly, the IEM method not only demonstrates the presence of DN within a cardiac cycle in its non-stationary component when DN is clearly visible but also reveals the location of DN within a cardiac cycle even when it is not visibly present in the input signal. The vertical dashed lines in Fig. 5 and Fig. 6 display the DN positions within a cardiac cycle estimated by the IEM-based algorithm. Moreover, Figs 7 (i-a) and 7 (i-b) demonstrate strong correlation for both ABP (*R*^2^(87343) =.99, *p*<.001) and PPG (*R*^2^(86764) =.98, *p*<.001) waveforms. *R*^2^ values close to 1 for both the ABP and PPG waveforms indicate a strong linear association between the SPD estimate by the algorithm and that marked by researcher as a reference.

**Fig. 5.**
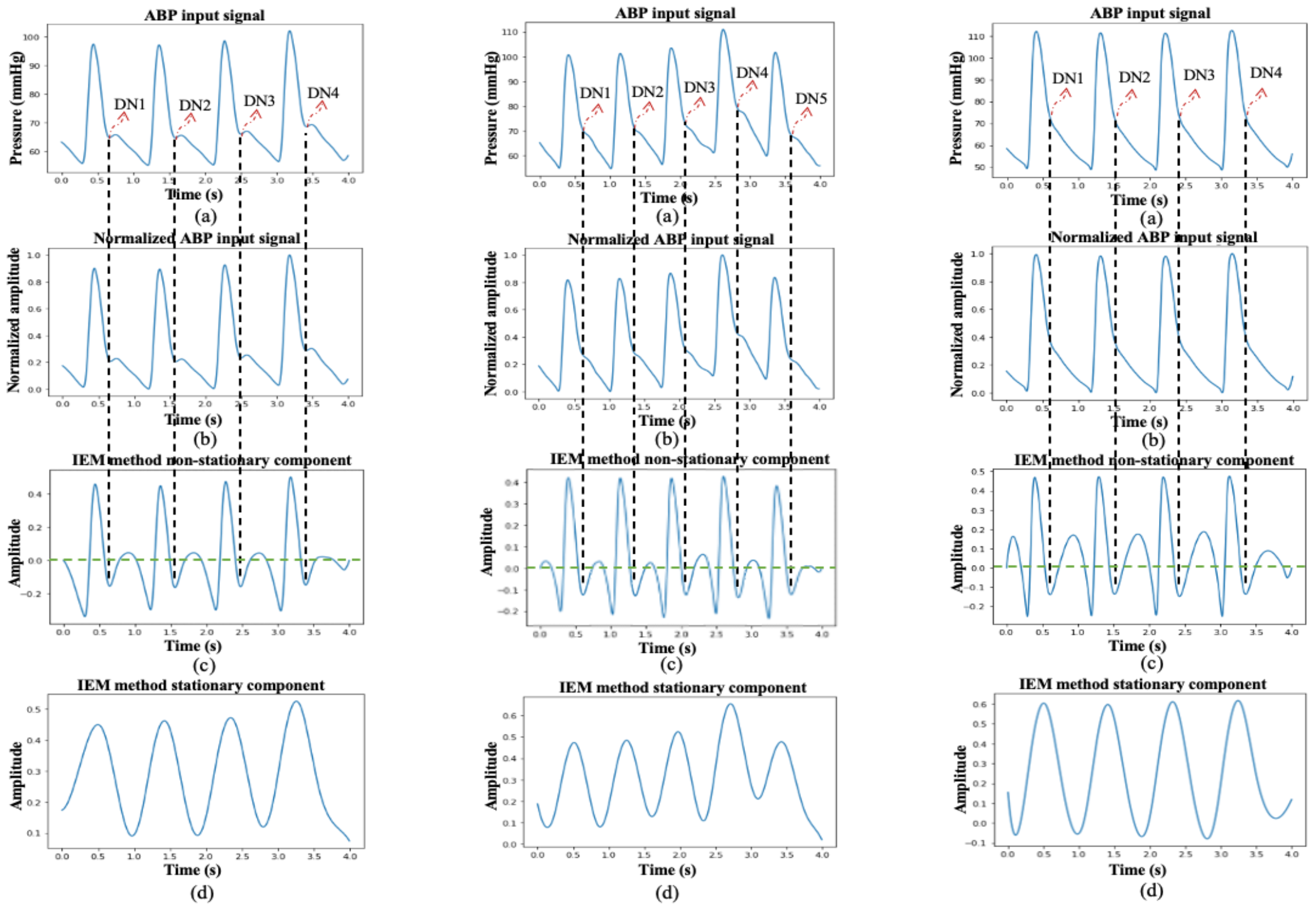
Three different cases of the process of DN detection using the IEM algorithm in ABP input signal: (i) where the DN is clearly observable within a cardiac cycle of ABP input signal; (ii) where DN is less evident within a cardiac cycle of ABP input signal; and (iii) where DN is absent within a cardiac cycle of ABP input signal. Plots labeled (a) depict the curves for 4-s ABP input signal window with marked DN locations by an experienced researcher in cases i and ii and with marked DN locations revealed by the IEM method in case iii, plots labeled as (b) depict the curves for the normalized input signal, plots labeled as (c) show the curves for the non-stationary component of the IEM method, and plots labeled as (d) display the curves for the stationary component of the IEM method.

**Fig. 6.**
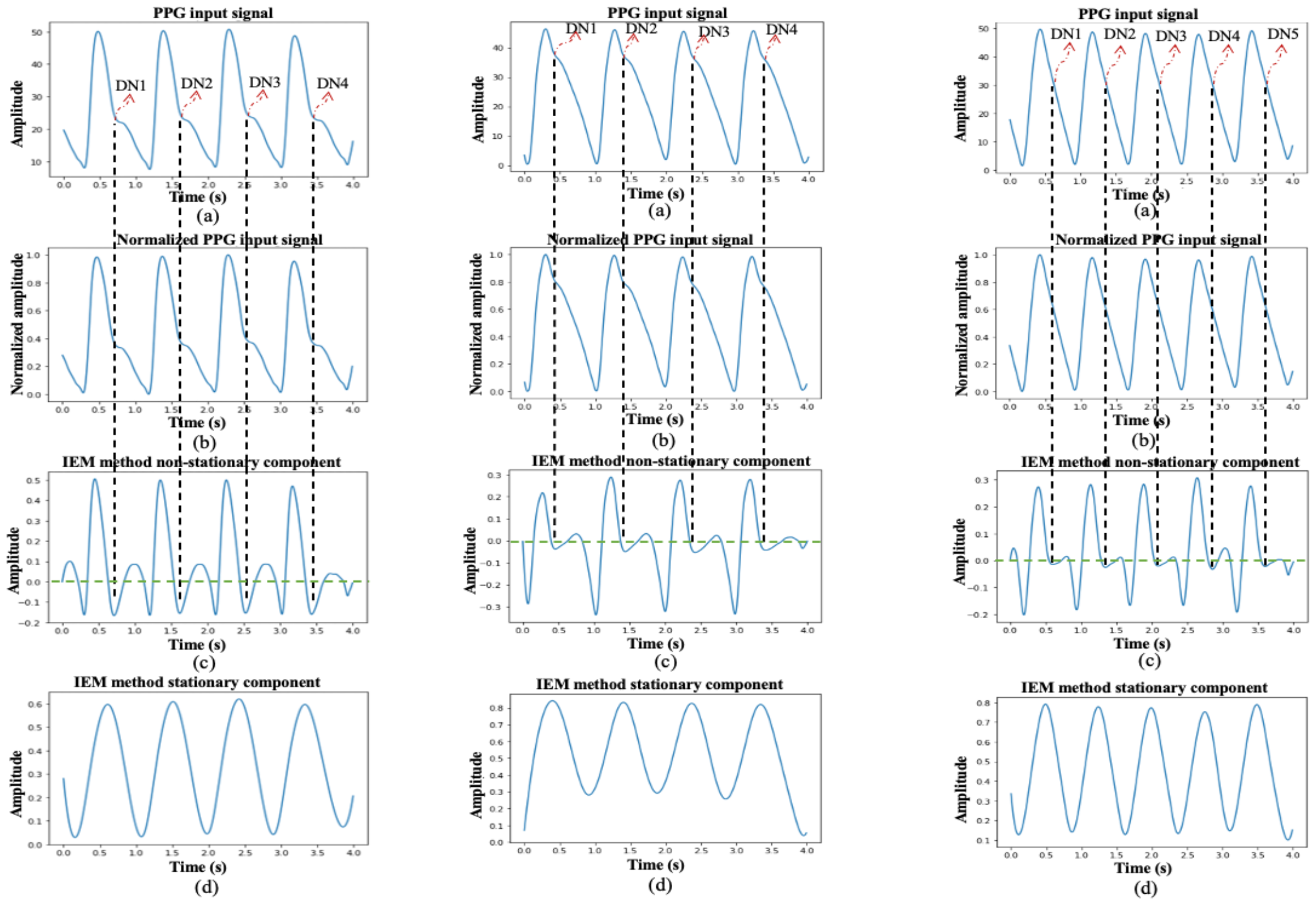
Three different cases of the process of DN detection using the IEM algorithm in PPG input signal: (i) where the DN is clearly observable within a cardiac cycle of PPG input signal; (ii) where DN is less evident within a cardiac cycle of PPG input signal; and (iii) where DN is absent within a cardiac cycle of PPG input signal. Plots labeled (a) depict the curves for 4-s PPG input signal window with marked DN locations by an experienced researcher in cases i and ii and with marked DN locations revealed by the IEM method in case iii, plots labeled as (b) depict the curves for the normalized input signal, plots labeled as (c) show the curves for the non-stationary component of the IEM method, and plots labeled as (d) display the curves for the stationary component of the IEM method.

**Fig. 7.**
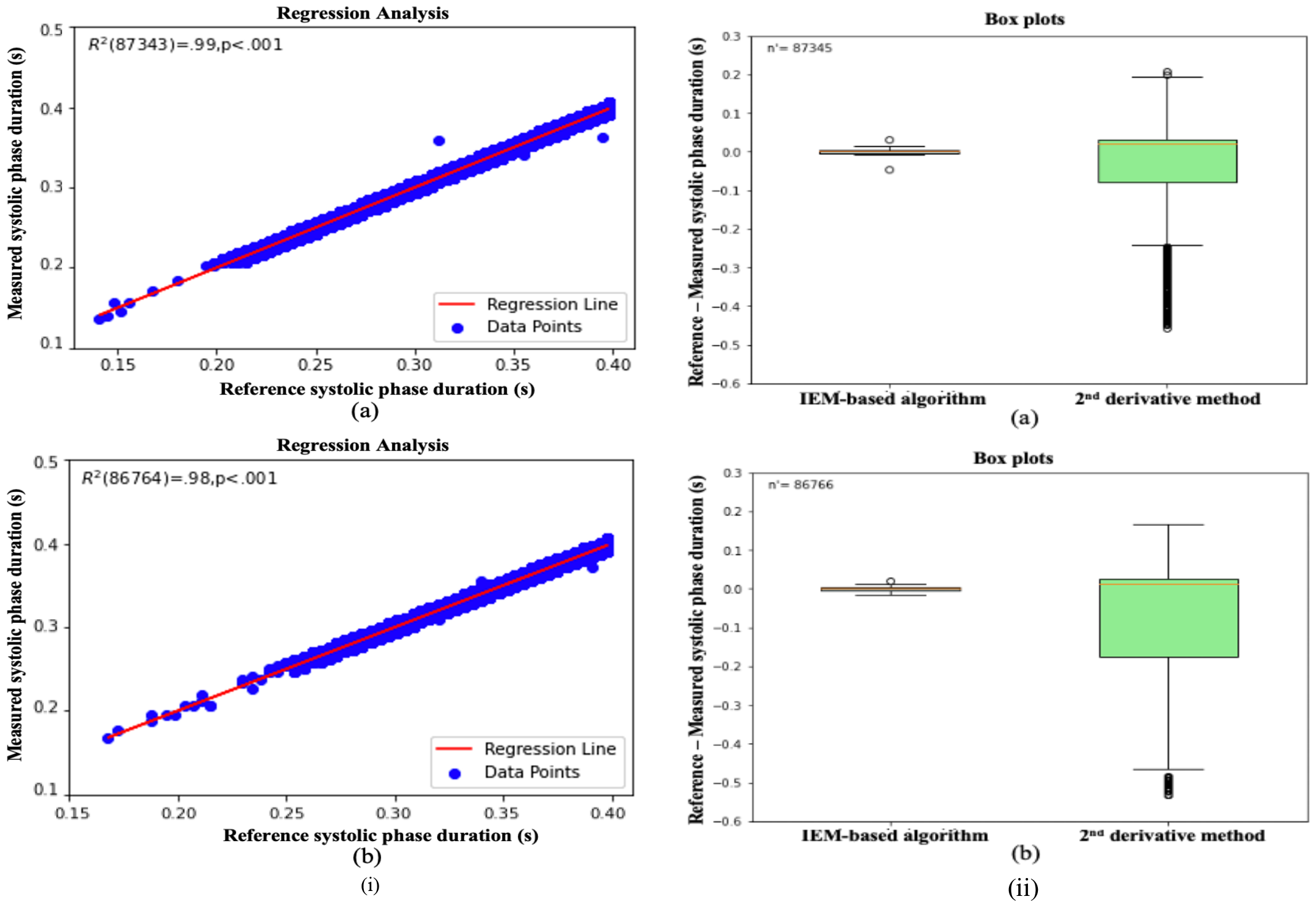
(i) (a) Regression analysis for the ABP waveforms, for the IEM method estimated systolic phase duration; (b) Regression analysis for the PPG waveforms, for the IEM method estimated systolic phase duration. (ii) Box Plots, comparison with the 2^nd^ derivative for ABP waveforms (a) and PPG waveforms (b).

### 4.2 Comparison of the IEM based algorithm with the 2^nd^ derivative method

The DN detection ability of the IEM-based algorithm was compared with the 2^nd^ derivative method using the rate of detectability (*D*_*R*_) and the difference in SPD (reference-measured), as shown by box plots (Fig. 7 (ii)). As presented in Table 1, both methods achieved a 100 % *D*_R_ for both ABP and PPG cardiac cycles, but the IEM-based algorithm gives, on average, estimates for the SPD which are closer to the reference estimates than the 2^nd^ derivative method. As mentioned earlier, the absolute value of the difference in SPD can be referred to as the error of DN detection. To assess the error on comparing it to three different permitted error ranges: 30 ms (the strict permitted error range), 50 ms (typical permitted error range), and 70 ms (tolerant permitted error range) [30], the IEM-based algorithm achieved an average error well below the strict permitted error range (30 ms) in both ABP (4.7 ms) and PPG (4.6 ms) cases. However, in the 2^nd^ derivative method, the average error was slightly below the tolerant permitted error range (70 ms) for ABP cases (69.3 ms), but for PPG cases (96.8 ms), it even exceeded the tolerant permitted error range (70 ms) significantly (see Table 1).

**Table 1:** Evaluation of DN detection of the IEM based algorithm in ABP and PPG cardiac cycles.

| Cases | $N_R$ | $\overline{SPD}_R$ (SD)<br>(s) | IEM based algorithm | | | | 2<sup>nd</sup> derivative method | | | |
| --- | --- | --- | --- | --- | --- | --- | --- | --- | --- | --- |
| | | | $N_D$ | $D_R$<br>(%) | $\overline{SPD}_M$ (SD)<br>(s) | $\overline{error}_{DN}$ (SD)<br>(s) | $N_D$ | $D_R$<br>(%) | $\overline{SPD}_M$ (SD)<br>(s) | $\overline{error}_{DN}$ (SD)<br>(s) |
| ABP ($n'=87,345$) | 87,345 | 0.3448<br>(0.0336) | 87,345 | 100 % | 0.3448<br>(0.0333) | 0.0047 (0.0029) | 87,345 | 100 % | 0.3675<br>(0.1005) | 0.0693 (0.0770) |
| PPG ($n'=86,766$) | 86,766 | 0.3564<br>(0.0286) | 86,766 | 100 % | 0.3565<br>(0.0283) | 0.0046(0.0029) | 86,766 | 100 % | 0.4212<br>(0.1154) | 0.0968 (0.0909) |

ABP: Arterial blood pressure; PPG: Photoplethysmography; $n'$ : Number of input cardiac cycles; $\overline{SPD}_R$ : Mean of reference systolic phase duration; SD: standard deviation; $\overline{SPD}_M$ : Mean of measured systolic phase duration by an algorithm; $\overline{error}_{DN}$ : mean error of DN detection; $N_R$ : Reference diastolic notches marked by an experienced researcher; $N_D$ : Detected diastolic notches by an algorithm; $D_R$ : Rate of detectability.

### 4.3 Robustness analysis of the IEM based algorithm

To assess the IEM-based algorithm’s robustness when the DN is not visibly distinct, we conducted a test across an SNR range from -30 dB to -5 dB. Fig. 8(i) and Fig. 8(ii) show the robustness performance of the proposed algorithm for ABP (*n*^′^ = 1,171,288) and PPG (*n*^′^ = 3,424,975) cardiac cycles, respectively, in terms of two quantitative evaluators: rate of detectability, and error of DN detection at the given SNR. Figures 8 (i-a) and 8 (ii-a) display curves for the average rate of detectability of ABP and PPG waveforms, respectively, across all cardiac cycles against SNR. Plots labeled (b) display the averaged error of DN detection over all cardiac cycles against SNR. We defined robust performance of the algorithm when the DN was not visibly distinct using the criterion: *D*_*R*_ ≥ 0.8, and error range of ≤ 0.045 s or 45 ms. For ABP cardiac cycles, the algorithm demonstrated robust performance for SNR values greater than or equal to -9 dB, while for PPG cardiac cycles, the algorithm indicated robust performance for SNR values greater than or equal to -12 dB.

**Fig. 8.**
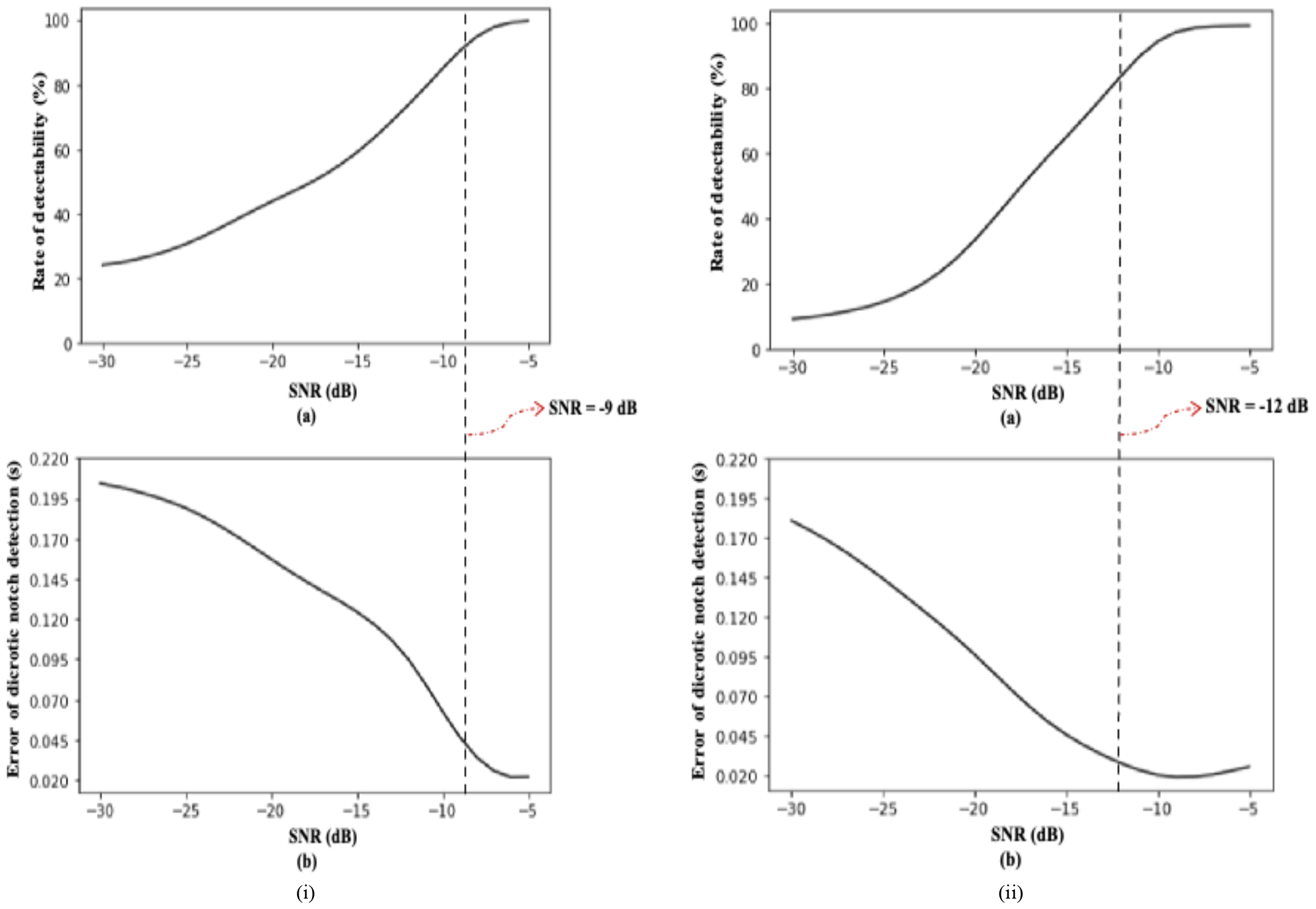
(i) (a) Rate of detectability plots for ABP waveforms with a signal to noise ratio in the range of -30 dB to -5 dB; (b) Systolic phase duration difference plots for ABP waveforms with a signal to noise ratio in the range of -30 dB to -5 dB. (ii) (a) Rate of detectability plots for PPG waveforms with a signal to noise ratio in the range of -30 dB to -5 dB; (b) Systolic phase duration difference plots for PPG waveforms with a signal to noise ratio in the range of -30 dB to -5 dB.

To further illustrate the effectiveness of the proposed algorithm for detecting indistinct DNs, we present one example at the threshold SNR value: ABP with SNR = -9 dB in Fig. 9 (i), and PPG with SNR = -12 dB in Fig. 9 (ii). Figs. 9 (a) display the 4-s window of the normalized input signals before scaling. The nonstationary component after applying the IEM method is shown in Figs. 9 (b). Figs 9 (c) display the normalized input signals after scaling at SNR=-9 dB for ABP and at SNR=-12 dB for PPG. The nonstationary component after applying the IEM method on the scaled and normalized input signals is displayed in Figs. 9 (d). Comparing the plots, we can see that the DN, clearly identifiable before scaling is less distinct afterwards for both ABP and PPG cardiac cycles. However, in both cases the IEM-based algorithm locates the DN in the same position.

**Fig. 9.**
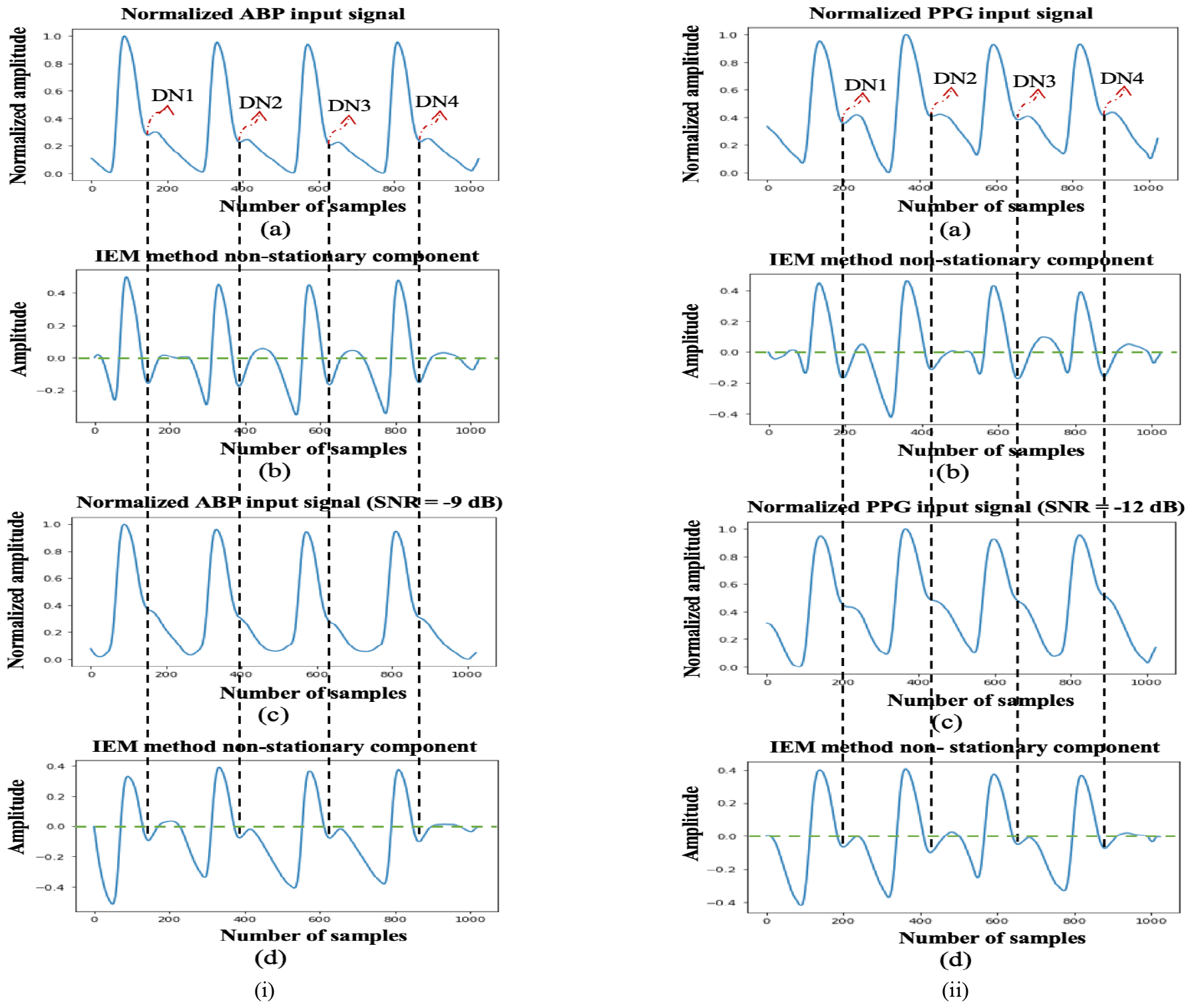
(i) (a) Normalized ABP input signal with marked dicrotic notch locations by an experienced researcher; (b) IEM method non-stationary component; (c) Normalized ABP input signal at SNR = -9 dB; (d) IEM method non-stationary component. (ii) ((a) Normalized PPG input signal with manually marked dicrotic notches; (b) IEM method non-stationary component; (c) Normalized PPG input signal at SNR = -12 dB; (d) IEM method non-stationary component.

## 5. Discussion

We have proposed a novel IEM-based algorithm for detecting the DN in ABP and PPG waveforms. The DN appears within the cardiac cycle, situated between the systolic peak and the diastolic endpoint, marking the transition from the systolic phase to the diastolic phase. Accurate detection of the DN in these waveforms is a crucial aspect of cardiovascular monitoring and clinical research. For example, as discussed in [9], ABP waveforms contain a range of information, including morphological features such as the position of the DN, which can provide valuable insights into overall hemodynamic status and may prove beneficial in managing complex hemodynamic situations. Furthermore, Abushouk et al. [4] mentioned that changes in DN parameters often appear early in the progression of vascular disease, potentially aiding in its early recognition. He et al. [41] noted that the DN in PPG waveforms may be useful for estimating blood pressure. Hermeling et al. [42] utilized the DN as a temporal reference point to measure arterial stiffness, which is essential for assessing cardiovascular risk. However, as shown in [23, 43], the DN tends to diminish with advancing age, which can make it challenging to locate in measurements from older subjects. Therefore, it is not only important to detect the DN when it is prominent within a cardiac cycle, but also in DN-less cardiac cycle [30].

The IEM-based algorithm has demonstrated the ability to locate the DN in both DN-present and DN-less ABP and PPG waveforms with a high rate of detectability and good robustness above SNRs of -9 dB for ABP and -12 dB for PPG waveforms. Moreover, in terms of computational cost, the IEM method requires only *O* (*IN*) operations for number of iterations *I* and signal length of *N*. This suggests that the IEM-based algorithm could serve as a trustworthy signal processing approach for precise DN localization and has the potential to support the clinical exploitation of DN as a valuable diagnostic and prognostic marker, ultimately enhancing patient care and medical research. In comparison to the existing 2^nd^ derivative method [32], where the reference for both methods was the DNs marked by an experienced researcher, the IEM-based algorithm offers an equally high rate of detectability but provides a closer estimate of SPD and lower error of DN detection in both ABP and PPG waveforms.

Our study has several limitations. First, the SG filter parameters in the IEM method are not adaptive. This lack of adaptability can be problematic, particularly in cases with high-frequency background noise, as mentioned in [1]. In such situations, it may lead to inappropriate estimation of the envelope mean value. Consequently, this can make it difficult for the IEM-based algorithm to accurately locate the DN in the non-stationary output of the IEM method; second, the dependency of the IEM method stopping criteria on the non-adaptive accuracy level (*β*), may affect whether the DN location is accurately identified in the non-stationary component of the IEM output; third, the algorithm depends on two empirically set conditions for assigning the valley corresponding to the DN location in the non-stationary component of the IEM method. This may affect whether the correct valley is assigned as the DN location.

## 6. Conclusions

This study presents the IEM-based algorithm for the detection of DN in both DN-present and DN-less ABP and PPG waveforms. The IEM-based algorithm was evaluated using a large perioperative dataset (MLORD) and compared with the existing 2^nd^ derivative method. The key findings of the study are: (1) The IEM-based algorithm has low computational cost, and demonstrates high accuracy in DN detection, achieving 100 % accuracy in both ABP and PPG waveforms; (2) The IEM-based algorithm can locate the DN even when it is not visibly identifiable in the input signal; (3) The IEM-based algorithm can determine the temporal location of DN within a cardiac cycle with lower error in DN detection in both ABP and PPG waveforms compared to the 2^nd^ derivative method.

We conclude that the IEM-based algorithm is suitable to use in a clinical context for accurately locating DN within a cardiac cycle. This information can be used to estimate time-domain features related to DN in both ABP and PPG waveforms, which in turn can contribute to cardiovascular research and clinical studies. Ultimately, this could enhance patient care and inform treatment decisions. Future research will focus on evaluating the performance of the IEM-based algorithm on a more diverse dataset collected from multiple centers that will further test its generalizability and real-world applicability.

## Data Availability

The interested parties may reach out to the corresponding author of this article to request the access to the MLORD dataset.

## Acknowledgments

This work was supported by the National Institutes of Health (NIH): R01EB029751 and R01HL144692.

